# Long-term Outcomes of Poor-grade Aneurysmal Subarachnoid Haemorrhage: A Multicentre Observational Cohort Study

**DOI:** 10.1101/2024.11.25.24317947

**Authors:** Bangyue Wang, Tianxing Li, Yan Zhao, Tian Zhou, Ruyi Wang, Yang Li, Xiuhu An, Jiheng Hao, Kaijie Wang, Xinyu Yang

**Affiliations:** Department of Neurosurgery, Tianjin Medical University General Hospital, Tianjin, 300070, China; Department of Neurosurgery, The Second Affiliated Hospital of Anhui Medical University, Hefei, 230601, China; Department of Neurosurgery, Affiliated Jinhua Hospital, Zhejiang University School of Medicine, Jinhua, 321000, Zhejiang, China; Department of Emergency Surgery, The First Affiliated Hospital of Bengbu Medical University, Bengbu, 233030, Anhui, China; Department of Neurosurgery, Liaocheng People’s hospital, Liaocheng, 252000, Shandong, China; Department of Neurosurgery, Tangshan Gongren Hospital, Tangshan, 063000, Hebei, China

**Keywords:** aneurysm, subarachnoid haemorrhage, outcome, poor-grade aSAH, survival analysis

## Abstract

**BACKGROUND:** Despite advancements in neurosurgery and intensive care that reduce overall mortality, poor-grade aneurysmal subarachnoid haemorrhage (aSAH) (World Federation of Neurosurgical Societies [WFNS] grades IV and V) remains a significant clinical challenge and is associated with persistently high mortality rates. The aim of this study was to assess the long-term outcomes of poor-grade aSAH and to identify factors influencing patient prognosis to guide clinical management.

**METHODS:** A multicentre, observational cohort study was conducted across 12 regional centres in northern China. The study included patients with poor-grade aSAH admitted from 2017 to 2020. The baseline data included demographics, clinical presentation, aneurysm characteristics, and treatment modalities. Outcome data, including survival status, mortality along with its associated causes and timing, and modified Rankin scale (mRS) scores, were collected prospectively at the last medical follow-up. Changes in case fatality over time were quantified with weighted linear regression. Survival analysis was performed to estimate survival and hazard ratios for death. Binary logistic regression was performed to estimate the odds ratio for dependency (mRS=3-5).

**RESULTS:** Among the 1,589 enrolled patients, 1,339 were successfully followed, with an average follow-up of 26.37 months. Among them, 61.5% (824/1,339) were dependent or died. The overall mortality rate was 51% (684/1,339), and 21.3% (140/655) of the survivors were dependent. The risk factors for mortality included age ≥65 years, previous history of stroke, and WFNS grade V. Additionally, conservative treatment and endovascular treatment were identified as risk factors and protective factors, respectively, compared with surgical treatment. WFNS grade V and middle cerebral artery aneurysms are independent risk factors for dependency.

**CONCLUSIONS:** Although there has been a downward trend in recent years, the long-term mortality rate for poor-grade aSAH has remained significantly high at 51%, with 21.3% of survivors being dependent. Active aneurysm treatment, to the extent possible, is crucial for improving the prognosis of these patients.

## INTRODUCTION

The mortality and disability rates associated with aneurysmal subarachnoid haemorrhage (aSAH) are exceedingly high, with the mortality rate reaching approximately 35%^1^. However, with the progress in modern neurosurgical techniques and the proliferation of neurointensive care units, there has been a significant reduction in the overall mortality rate of aSAH^2–6^. Despite this encouraging trend, the mortality rate for poor-grade (World Federation of Neurosurgical Societies [WFNS] grade IV and V) aSAH patients remains high. Although poor-grade aSAH accounts for only approximately 18–30% of all aSAH cases^7–9^, it contributes to more than half of aSAH-related deaths^10–12^, and the management of these patients remains a clinical challenge. Previous studies have demonstrated that more than 60% of these patients die or are left with disabilities^9, 13, 14^, imposing a significant burden on society and families^15, 16^. Therefore, describing the long-term prognosis of poor-grade cases and analysing their influencing factors are crucial for guiding the management of patients with poor-grade aSAH in clinical practice and preventing the occurrence of adverse events in the long term. To the best of our knowledge, current reports on the long-term prognosis of poor-grade aSAH are mostly based on small sample sizes and are single-centre retrospective studies^17^, which are not able to fully reflect the epidemiological characteristics and long-term prognosis of the poor-grade aSAH population. In this study, we used the Chinese Multicentre Cerebral Aneurysm Database to analyse long-term outcomes and additional survival data of poor-grade aSAH patients to provide clinical evidence that could help enhance the understanding of patient prognosis and assist in providing informed and reasonable management for this challenging subgroup.

## METHODS

### Patients

This prospective, multicentre, observational cohort study (chictr.org.cn, ID: ChiCTR2200065083) was approved by the Ethics Committee (the Ethics Committee of Tianjin Medical University General Hospital (IRB2021-YX-178-01)) and was performed in accordance with the Declaration of Helsinki. The requirement for informed consent was waived by the ethics committee because of the observational design of this study.

This study utilised the Chinese Multicentre Cerebral Aneurysm Database, which consecutively enrolled patients from 12 regional centres across 4 provinces in northern China. Data from all patients with aneurysmal subarachnoid haemorrhage (SAH) admitted from 2017 to 2020 were entered into the Chinese Multicentre Aneurysm Database (CMAD).

The inclusion criteria for the present study were all consecutive adult patients (≥18 years) admitted to the neuro-intensive care unit (ICU) or neurosurgical department because of the first diagnosis of poor-grade SAH, defined as WFNS grades IV and V with an aneurysmal cause. The noninclusion criteria were aSAH rehospitalization, poor echogenicity or inability to obtain inpatient medical records, baseline characteristics or telephone number. Among the 5837 aSAH patients in the database, 1589 eligible patients admitted with WFNS grades IV and V were identified.

### Study design and data collection

For eligible patients included in the study, baseline demographic data, WFNS grade, and Fisher grading scale score were directly obtained by neurosurgeons at each centre upon admission and recorded from admission to discharge. In addition, the size of the aneurysm, aneurysm multiplicity, aneurysm location, and aneurysm treatment methods were recorded. The diagnosis of SAH was confirmed by head computed tomography (CT) or lumbar puncture. Evidence of aneurysms was verified by neuroradiologists through computed tomography angiography (CTA), digital subtraction angiography (DSA), or magnetic resonance angiography (MRA). The clinical management of patients and the indications for different aneurysm treatment strategies were determined by an interdisciplinary neurovascular team on the basis of aneurysm treatment guidelines after discussion with family members. The final treatment methods included surgical treatment, endovascular treatment, or conservative treatment. Conservative treatment was defined as not performing surgical repair for ruptured aneurysms. In addition to medication, these patients may have undergone simple external drainage surgery, simple haematoma removal, or decompressive craniectomy, which are interventions taken for patients with gradually declining consciousness due to hydrocephalus or intracerebral haemorrhage (ICH).

### Outcome measures

Survival analysis data were collected starting on the day of ictus. The follow-up evaluation was accomplished primarily via standardized telephone surveys with the patients or close relatives. The final follow-up period was from June 1, 2023, to June 1, 2024. The outcome measures included survival and mortality status at discharge and at the time of follow-up, as well as the modified Rankin scale (mRS) score. Additionally, the primary outcome was death, and the secondary outcome was dependent living status (dependency), defined as an mRS score of 3–5 (indicating an inability to walk unassisted) at the time of follow-up. The patient characteristics analysed included sex, age, living area (rural/urban), hypertension, diabetes, hyperlipidaemia, smoking, alcohol consumption, previous history of stroke, aneurysm multiplicity, aneurysm location, WFNS grade, Fisher grading scale on admission, presence of ICH or intraventricular haemorrhage (IVH) on admission, aneurysm size, admission delay, intervention delay (surgical and endovascular treatment), length of hospital stay, cause of death, and general information on treatment (surgical, endovascular, or conservative treatment; use of lumbar puncture or external ventricular drainage). The causes of death are classified as follows: (1) direct effect of the primary haemorrhage, (2) aneurysm rebleeding, (3) refractory cerebral oedema leading to brainstem herniation, (4) withdrawal of life-sustaining treatment for patients in a persistent vegetative state, (5) infection, (6) multiple organ dysfunction syndrome (MODS), (7) hydrocephalus, (8) cardiac disease, (9) other cerebrovascular disease (e.g., intracerebral haemorrhage, cerebral infarction), (10) cancer, (11) pulmonary embolism, (12) accidents (e.g., car accidents), and (13) unknown.

### Statistical methods

Continuous data are described using the mean (SD) or median (interquartile range), depending on their distribution. Categorical data are described as counts (percentages). Multiple imputation methods are used to address missing data issues. The fully conditional specification method is employed to estimate missing values of covariates via iterative Markov chain Monte Carlo methods. Kaplan‒Meier curves were plotted for survival analysis of different groups and the overall population. Cox proportional hazard regression models were used to estimate the hazard ratio (HR) of death after poor-grade aSAH. Binary logistic regression models were used to identify factors impacting dependent living (defined as mRS 3-5) after poor-grade aSAH. The variables for multivariate analysis included sex, age, and variables with p<0.2 in the univariate analysis. The p value and 95% confidence interval (CI) were calculated. On this basis, the cumulative mortality rates at various time points for the overall risk factor and each risk factor are summarized. Linear regression, weighted by the inverse of the standard error of the case fatality rate for each study, was used to quantify the relationship between the case fatality rate and the mid-central year of the study. Continuous variables were tested via t tests or Wilcoxon tests as appropriate. Pearson’s chi-square analysis was used to test for differences in the stratified distributions of various factors. All analyses were conducted via SPSS version 27.0 (SPSS Inc., Chicago, IL) or R version 4.0.3(https://www.R-project.org/).

## RESULTS

### Baseline characteristics

From 2017–2020, a total of 9,826 eligible intracranial aneurysm patients were included, 5,837 of whom with subarachnoid haemorrhage (SAH) met the criteria, and a total of 1,589 eligible consecutive poor-grade aSAH patients were included (Figure S1). In our study, they accounted for 27.2% of the overall SAH cases. Among the 1,589 poor-grade aSAH patients included in this study, the average age (SD) of the patients was 62.31 years (11.57), with 67.7% being female. There were 1,005 WFNS Grade IV patients and 584 WFNS Grade V patients. The location of the aneurysm was missing for 12 (0.08%) patients, with the most common location being the internal carotid artery (ICA, 39.5%). Data on the maximum diameter of the aneurysm were missing for 269 (20%) patients, with the most common size being 4–6.9 mm, totalling 344 (35.5%) patients. The treatments included surgical, endovascular, and conservative treatment, with 670 (42.1%) patients receiving surgical treatment, 539 (49.2%) patients receiving endovascular treatment, and 380 (23.9%) patients receiving conservative treatment (Table 1).

**Table 1.**
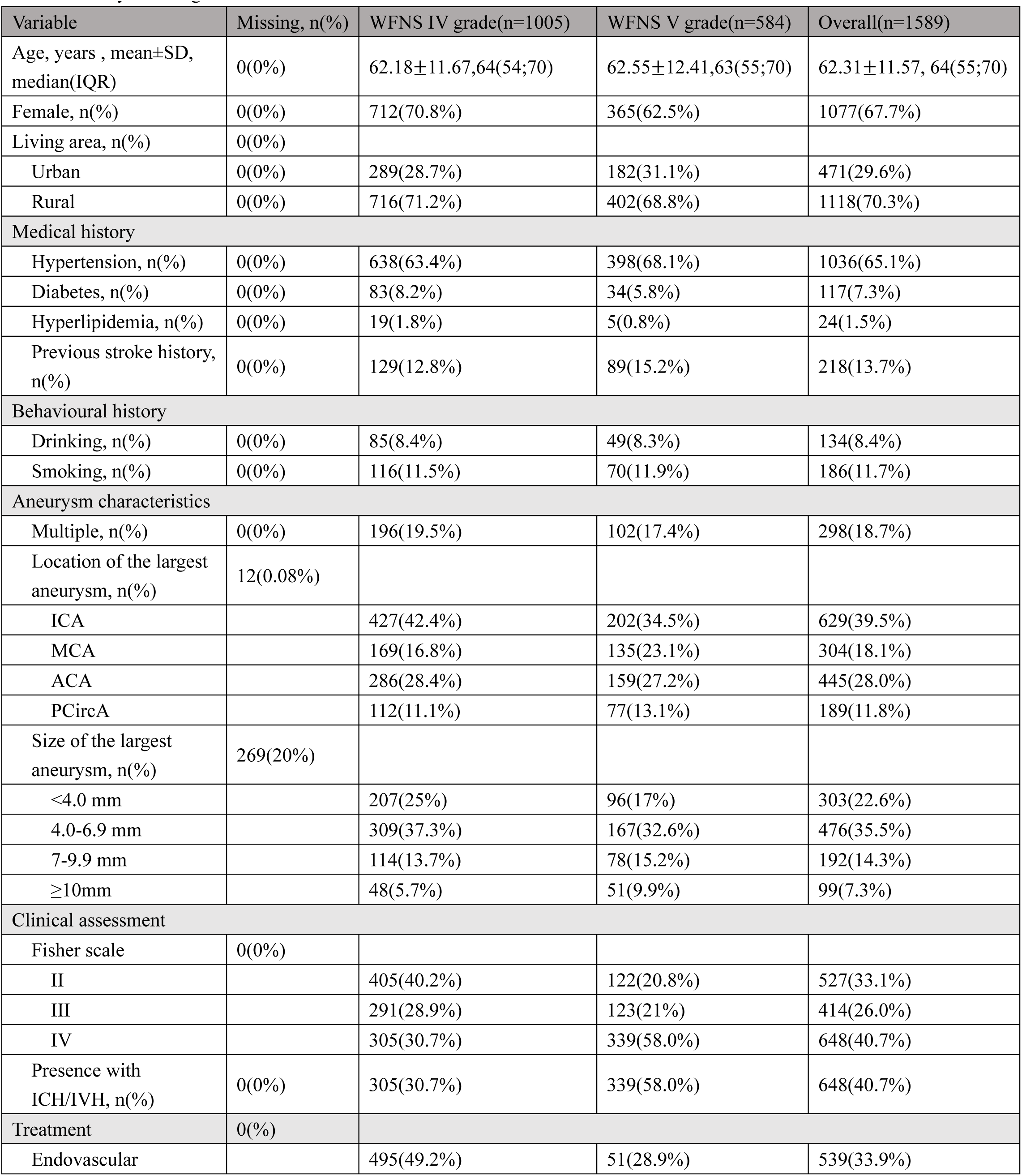
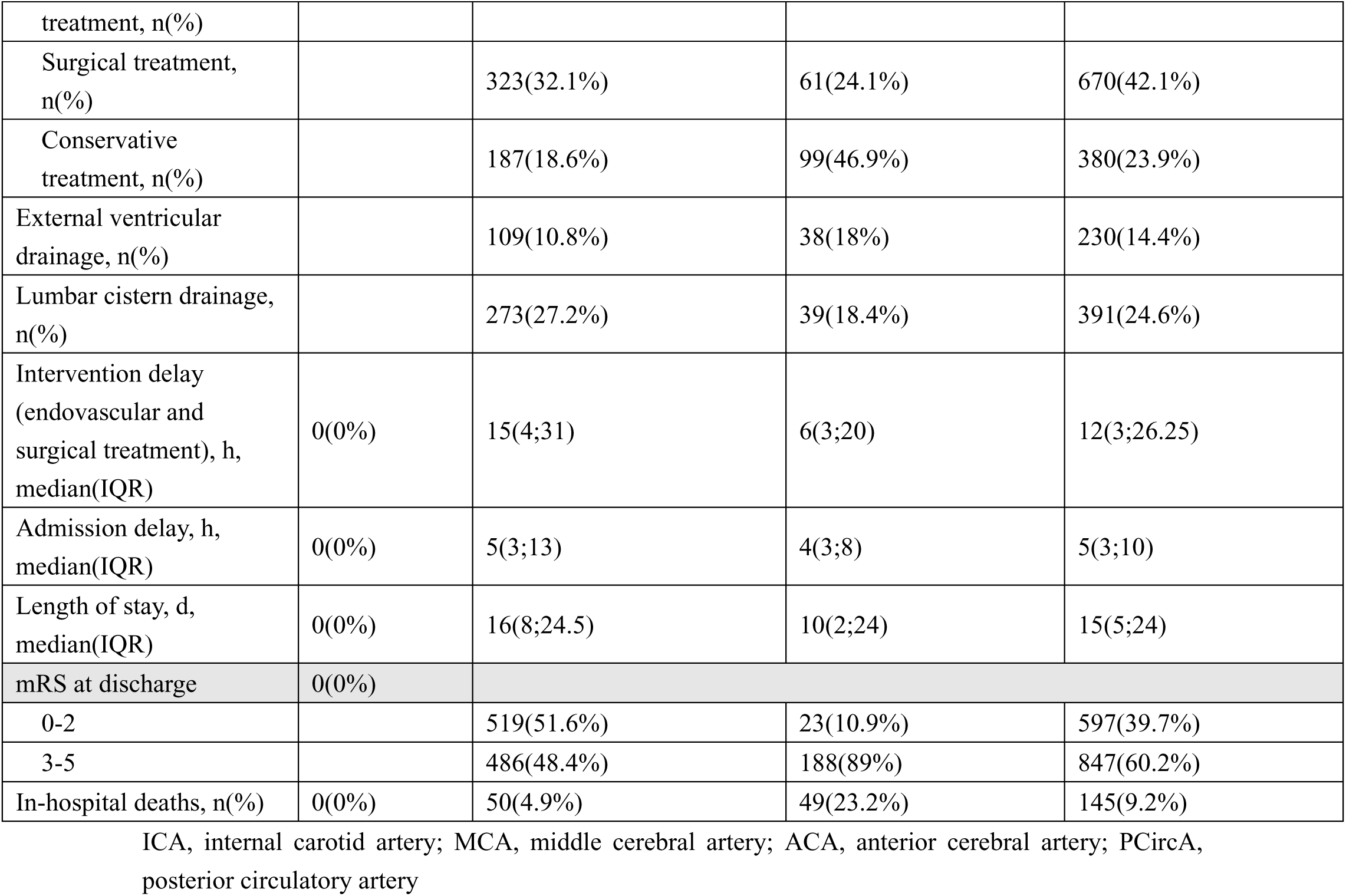
Baseline characteristics table of 1,589 poor-grade subarachnoid haemorrhage patients grouped by WFNS grade at admission.

### Survival analysis

In this study, we used the Kaplan‒Meier method to generate time‒survival-related survival curves on the basis of associated factors, as shown in Figure 1. A total of 1,339 out of 1,589 patients were included in the survival analysis, accounting for 2,942.83 follow-up years, with an average follow-up of 26.37 months (range, 0.5–88 months). During the follow-up, a total of 61.5% (824 out of 1,339) of the patients either died or survived with dependency status.

**Figure 1.**
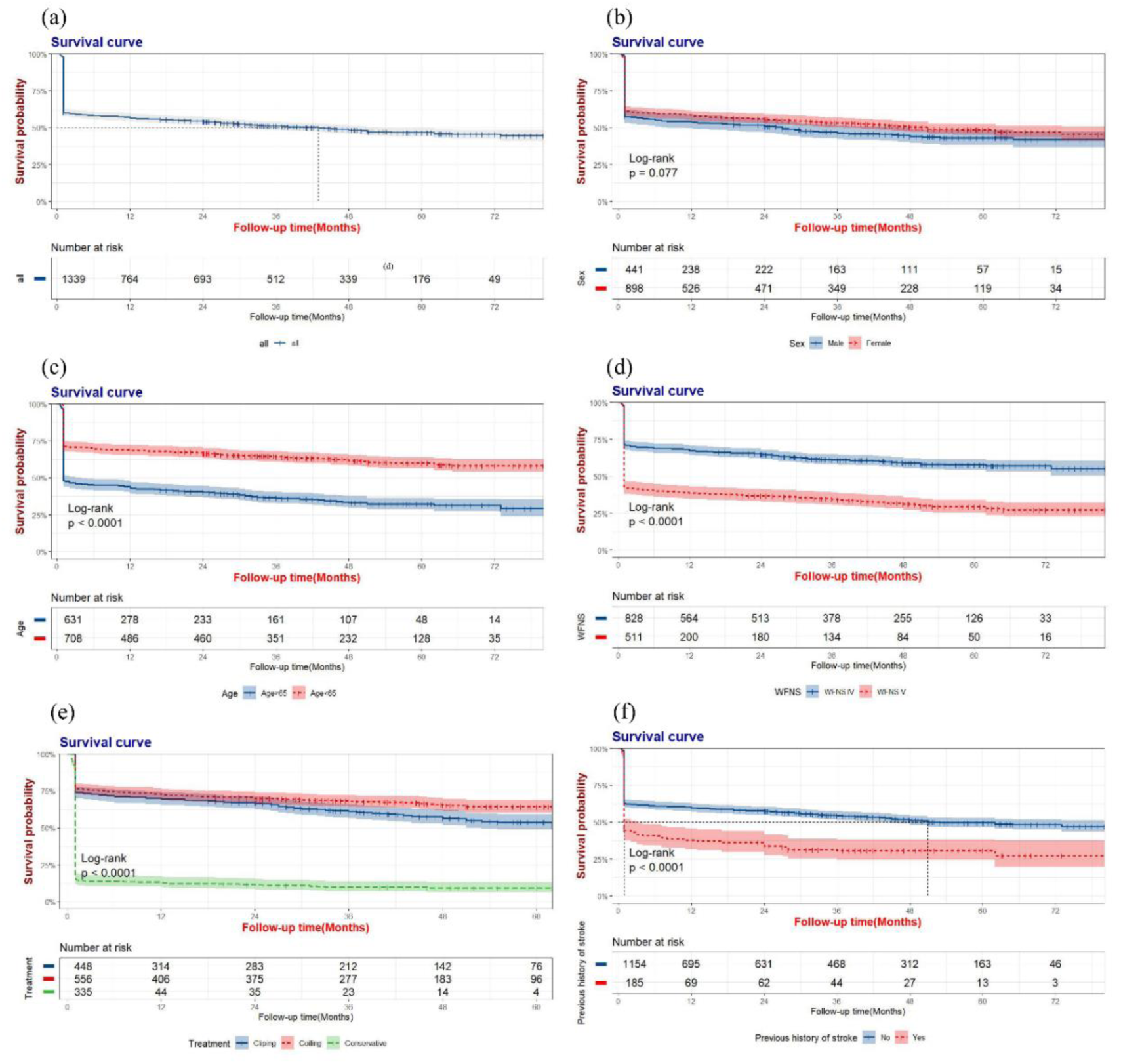
Probability of survival by risk factors.(a), Kaplan-Meier curves of the complete cohort, (b-f) stratified by sex (b),age (c),WFNS grade (d),treatment (e) and previous history of stroke (f).

There were 684 (51%) patients with poor-grade aSAH who died from all causes, while 655 (49%) patients were still alive at the time of follow-up. Figure S2 illustrates the distribution of causes of death, whereas Table S1 elaborates on their temporal distribution. The survival rates at various time points related to various influencing factors are shown in Table 2. Of these rates, 145 patients died in the hospital.

**Table 2.** Cumulative 3-year survival probability rate (95% CI) according to different associated factors.

|  | 1mo(95%CI) | 3mo(95%CI) | 6mo(95%CI) | 1y(95%CI) | 2y(95%CI) | 3y(95%CI) |
| --- | --- | --- | --- | --- | --- | --- |
| All | 60.3%<br>(57.8%-63.0%) | 59.1%<br>(56.5%-61.8%) | 58.0%<br>(55.4%-60.7%) | 56.5<br>(53.9%-59.2%) | 54.1%<br>(51.5%-56.8%) | 51.1%<br>(48.5%-53.9%) |
| Age≥60 | 53.6%<br>(50.3%-57.0%) | 51.8%<br>(48.6%-55.3%) | 50.8%<br>(47.5%-54.2%) | 48.9%<br>(45.6%-52.4%) | 46.1%<br>(42.9%-49.6%) | 42.7%<br>(39.5%-46.2%) |
| Female | 61.7%<br>(58.6%-65.0%) | 60.2%<br>(57.1%-63.5%) | 59.4%<br>(56.2%-62.7%) | 57.8%<br>(54.7%-61.1%) | 55.6%<br>(52.5%-59.0%) | 53.3%<br>(50.2%-56.7%) |
| Hypertension | 56.1%<br>(52.9%-59.5%) | 54.8%<br>(51.6%-58.2%) | 53.7%<br>(50.5%-57.1%) | 51.9%<br>(48.7%-55.3%) | 49.0%<br>(46.3%-53.0%) | 47.1%<br>(43.9%-50.5%) |
| Previous history of stroke | 44.9%<br>(38.2%-52.6%) | 41.6%<br>(35.0%-49.3%) | 40.5%<br>(34.0%-48.2%) | 37.8%<br>(31.4%-45.4%) | 33.8%<br>(27.6%-41.4%) | 30.6%<br>(24.5%-38.1%) |
| Presence of ICH/IVH | 54.6%<br>(50.5%-58.9%) | 53.30%<br>(49.3%-57.6%) | 51.80%<br>(47.8%-56.2%) | 50.50%<br>(46.5%-54.9%) | 48.10%<br>(44.1%-52.5%) | 45.10%<br>(41.1%-49.6%) |
| WFNS V grade | 42.5%<br>(38.4%-47.0%) | 41.5%<br>(37.4%-46.0%) | 40.1%<br>(36.1%-44.6%) | 38.7%<br>(34.7%-43.2%) | 36.8%<br>(32.8%-41.2%) | 34.6%<br>(30.7%-39.1%) |
| 7-9.9 mm | 53.6%<br>(47.4%-60.6%) | 52.7%<br>(46.5%-59.7%) | 52.3%<br>(46.1%-59.3%) | 50.0%<br>(43.8%-57.0%) | 47.6%<br>(41.4%-54.7%) | 44.6%<br>(38.4%-51.9%) |
| ≥10mm | 59.4%<br>(50.8%-69.6%) | 57.5%<br>(48.9%-67.8%) | 57.5%<br>(48.9%-67.8%) | 55.7%<br>(47.0%-66.0%) | 51.9%<br>(43.2%-62.3%) | 49.7%<br>(41.0%-60.3%) |
| Surgical treatment | 73.9%<br>(69.9%-78.1%) | 72.5%<br>(68.5%-76.8%) | 71.0%<br>(66.9%-75.3%) | 69.%<br>(65.5%-74.0%) | 66.70%<br>(62.5%-71.2%) | 61.0%<br>(56.6%-65.8%) |
| Endovascular treatment | 76.3%<br>(72.8%-79.9%) | 75.5%<br>(72.0%-79.2%) | 74.1%<br>(70.5%-77.8%) | 72.3%<br>(68.0%-76.1%) | 69.7%<br>(66.0%-73.7%) | 68.0%<br>(64.2%-72.0%) |
| Conservative treatment | 15.8%<br>(12.4%-20.3%) | 13.7%<br>(10.5%-18.0%) | 13.7%<br>(10.5%-18.0%) | 12.8%<br>(9.7%-17.0%) | 11.3%<br>(8.4%-15.3%) | 9.7%<br>(7.0%-13.6%) |

In the Cox regression analysis, age ≥65 years (HR (95% CI): 1.677 (1.423-1.976), p<0.001), previous history of stroke (HR (95% CI): 1.280 (1.051-1.557), p=0.014), WFNS grade V (HR (95% CI): 1.566 (1.330-1.845), p<0.001), and conservative treatment (HR (95% CI): 2.580 (2.123-3.136), p<0.001) were identified as risk factors for death in poor-grade aSAH patients, whereas being female (HR (95% CI): 0.822 (0.700-0.966), p=0.017) and the use of endovascular treatment compared with surgical treatment (HR (95% CI): 0.810 (0.661-0.994), p=0.044) were protective factors (Table 3).

**Table 3.** Univariate and multivariable cox proportional hazards analysis of factors associated with primary outcome.

| Variable | Univariate analysis |  | Multivariate analysis |  |
| --- | --- | --- | --- | --- |
|  | P-value | HR (95% CI) | P-value | HR (95% CI) |
| Age≥65 | <0.001 | 1.982(1.699-2.312) | <0.001 | 1.677(1.423-1.976) |
| Female | 0.135 | 0.887(0.759-1.038) | 0.017 | 0.822(0.700-0.966) |
| Living area (reference to rural) | 0.530 | 0.949(0.806-1.117) |  |  |
| Hypertension | <0.001 | 1.411(1.195-1.666) | 0.052 | 1.184(0.999-1.402) |
| Diabetes | 0.342 | 1.143(0.868-1.506) |  |  |
| Previous stroke history | <0.001 | 1.635(1.349-1.983) | 0.014 | 1.280(1.051-1.557) |
| Current alcohol use | 0.427 | 0.894(0.677-1.180) |  |  |
| Current smoker | 0.899 | 1.015(0.805-1.280) |  |  |
| Presence of ICH/IVH | <0.001 | 1.294(1.113-1.504) | 0.349 | 1.079(0.920-1.265) |
| WFNS V grade (reference to IV grade) | <0.001 | 1.930(1.659-2.244) | <0.001 | 1.566(1.330-1.845) |
| Multiple | 0.386 | 1.087(0.900-1.313) |  |  |
| Location of the largest aneurysm (reference to ICA) |  |  |  |  |
| MCA | 0.892 | 1.015(0.824-1.249) |  |  |
| ACA | 0.513 | 0.940(0.780-1.132) |  |  |
| PirCA | 0.211 | 1.162(0.918-1.470) |  |  |
| Size of the largest aneurysm (reference to <4 mm) |  |  |  |  |
| 4.0-6.9 mm | 0.377 | 1.086(0.905-1.303) | 0.186 | 1.131(0.942-1.359) |
| 7-9.9 mm | 0.044 | 1.263(1.006-1.586) | 0.780 | 1.033(0.821-1.300) |
| ≥10mm | 0.193 | 1.215(0.906-1.603) | 0.558 | 1.093(0.812-1.473) |
| Treatment (reference to surgical treatment) |  |  |  |  |
| Endovascular treatment | 0.010 | 0.768(0.628-0.939) | 0.044 | 0.810(0.661-0.994) |
| Conservative treatment | <0.001 | 3.124(2.593-3.764) | <0.001 | 2.589(2.131-3.146) |
| Admission delay, h | 0.196 | 1.001(1.000-1.001) | 0.218 | 1.001(1.000-1.001) |

ICA, internal carotid artery; MCA, middle cerebral artery; ACA, anterior cerebral artery; PCirCA,
posterior circulatory artery

### Assessment of functional outcomes

Information on outcomes was available for 1,589 patients at discharge and for 1,399 patients after follow-up. Among them, 36.8% (515/1399) had favourable outcomes (mRS 0-2), and 10.1% (140/1399) were dependent (mRS 3-5) (Table S2). The distribution of mRS scores is shown in Figure 2. Among the 655 patients who were alive, 78.6% (515) had favourable outcomes, and 21.3% (140) were dependent. Binary logistic regression analysis of the mRS score at the time of follow-up (Table S3, Figure S3) revealed that WFNS grade V (OR [95% CI]: 3.593 [2.354–5.484], p<0.001) and aneurysms in the middle cerebral artery (OR [95% CI]: 1.815 [1.053–3.127], P=0.032) were significantly associated with a greater risk of dependency.

**Figure 2.**
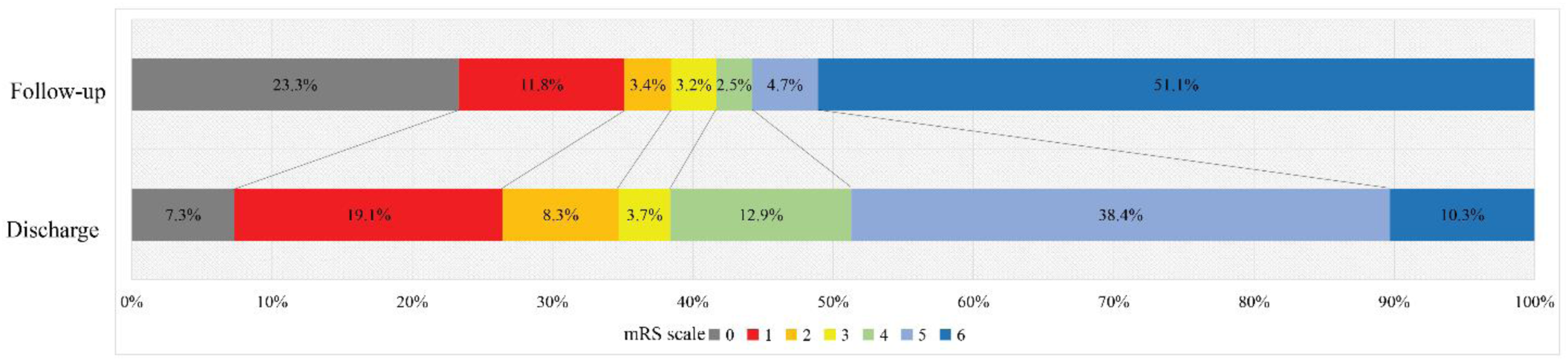
Distribution of modified Rankin Scale (mRS) scores at discharge and at the time of follow-up.

### Subgroup analysis

Figure 3 and Table S4 present subgroup analyses of the associations between the three types of treatment modalities and the risk of death. Compared with surgical treatment, conservative treatment was associated with a greater risk of death in all subgroups (P<0.05). Compared with surgical treatment, endovascular treatment was associated with significantly better outcomes in patients under 65 years of age and those who were male, nonhypertensive, without a previous history of stroke, without the coexistence of ICH/IVH, and with medium-sized aneurysms of 4–6.9 mm. Notably, the WFNS grade may not influence the choice between surgical and endovascular treatment (p>0.05). Interaction analysis indicated that conservative treatment may be safer for patients over 65 years old or males than for those under 65 years old or females.

**Figure 3.**
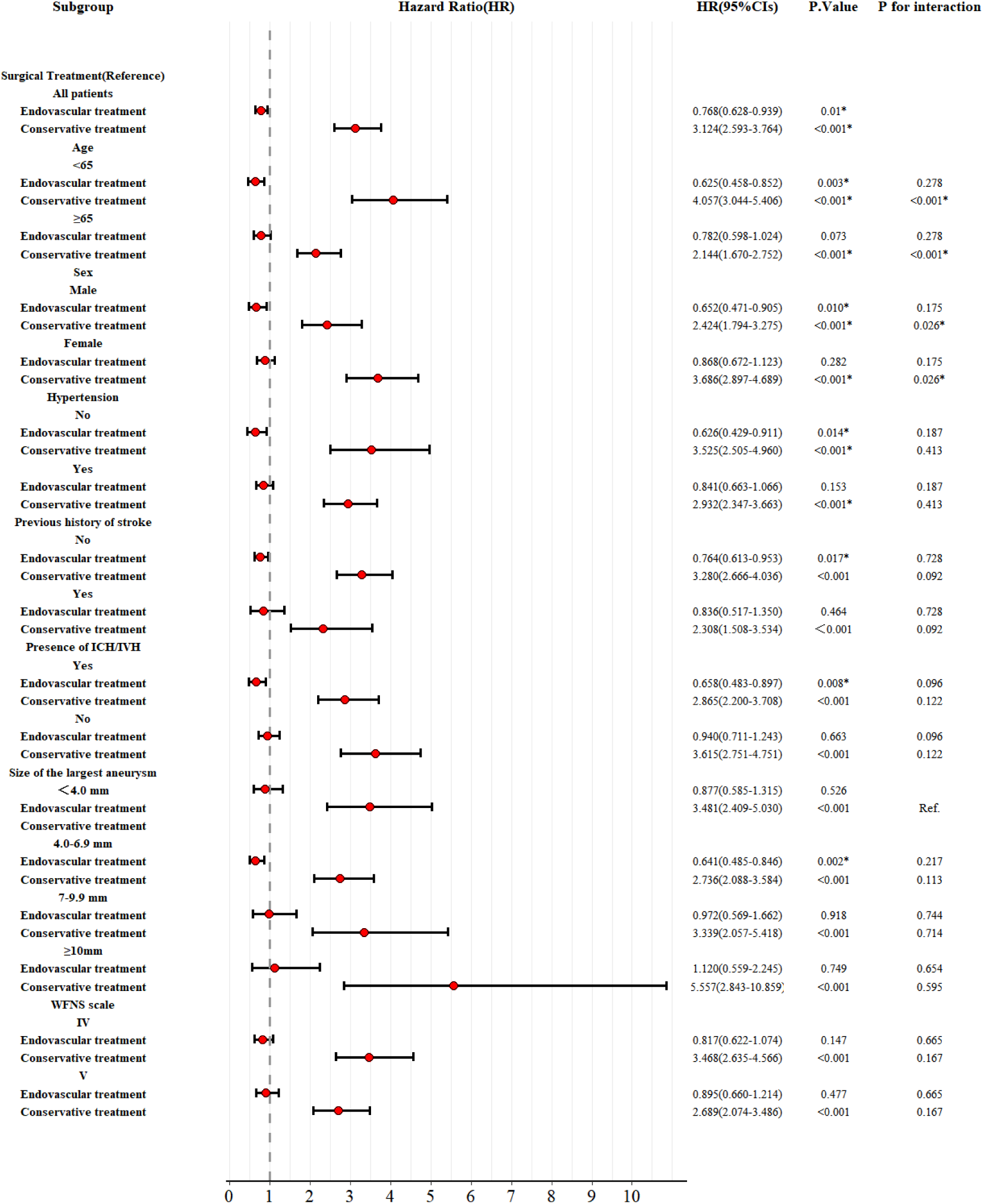
Subgroup analyses for the association between the treatment modality and death (surgical treatment is the reference for analysis)

## DISCUSSION

To our knowledge, the current analysis represents the largest prospective, multicentre registry of consecutive patients presenting with poor-grade aSAH. A total of 61.5% of the patients either died or became dependent on survival. Our results indicate that the long-term mortality rate for poor-grade aSAH in the Chinese population is 51%, and even among those who survived, 21.8% of the patients were left with dependency status. Additionally, we estimated the risk factors contributing to death and dependency. Our findings have important implications for evaluating the prognosis and management of poor-grade aSAH.

In this study, we observed that the long-term all-cause mortality rate for severe aSAH patients was 51.1%, with all-cause mortality rates at 3 months, 1 year, and 3 years being 40.9%, 43.5%, and 48.9%, respectively. de Oliveira Manoel retrospectively analysed 23 cohorts of unselected poor-grade SAH patients treated from 1977 to 2014 in 10 countries and reported that the mortality rate was approximately 60%^17^. This study reflects the mortality rate of poor-grade aSAH prior to 2014, which was higher than the mortality rate in our study, with patients admitted between 2017 and 2020. Furthermore, we performed a review of population-based case fatality after poor-grade aSAH to assess whether case fatality had changed. After excluding studies with evident selection bias and those with follow-up times of less than three months, a weighted linear regression analysis of the 19 studies (including the present study) evaluating mortality revealed that the case fatality rate decreased by 0·6% per year (95% CI 0·14 to 0·97), which, according to the model, corresponded to a 23% (6 to 40) decrease in case fatality over the 30 years studied (Table S5). This trend is consistent with the declining overall mortality rates of subarachnoid haemorrhage in recent years^2, 3^. Therefore, not only the overall mortality rate but also the mortality rate of poor-grade aSAH is believed to decrease over time; this is potentially due to early treatment (resulting from improved logistics and national health system coverage), recent improvements in neurocritical care, the widespread use of cerebrospinal fluid (CSF) drainage to buffer the massive increase in intracranial pressure (ICP) during rebleeding^18^, the application of continuous electroencephalography, invasive brain multimodality monitoring, and the use of nimodipine in the treatment of cerebral vasospasm^5, 12, 19^. Furthermore, advancements in aneurysm repair techniques, including the maturation of surgical treatment and the widespread use of endovascular treatment, have also led to some improvement in the mortality rate after poor-grade aSAH. Although the mortality rate has declined in recent years, it was relatively higher in the present study than in other countries in recent years, which may be related to a greater proportion of patients being treated conservatively. This phenomenon may reflect the particularity of China’s medical environment, including special medical policies and conservative treatment choices under family interventions^20^.

Survival analysis revealed that age≥65 years, previous history of stroke, WFNS grade V, and conservative treatment were risk factors for poor-grade aSAH. Age≥65 years, WFNS grade V, previous history of stroke, and conservative treatment as risk factors for poor-grade aSAH have been repeatedly mentioned in previous studies^21–23^. We found that endovascular treatment is a protective factor compared with surgical treatment, which is consistent with recent research findings^22^. Interestingly, female sex is a protective factor against poor-grade aSAH, possibly because of regional and hormonal factors^15^.

Through subgroup analysis of treatment methods, similar to previous studies, conservative treatment had a worse prognosis in all subgroups compared with surgical or endovascular treatment. In addition, in the subgroup analysis for treatment, we discovered that endovascular treatment is superior to surgical treatment in the subgroup of patients who are under 65 years old, male, nonhypertensive, without a history of stroke, without coexisting ICH/IVH, and with aneurysm diameters between 4–6.9 mm. Overall, endovascular treatment is better than surgical treatment and is now widely used in clinical practice with good outcomes. Interaction tests revealed that the disadvantage of conservative treatment is more pronounced in the younger group (aged <65) than in the older group (aged ≥65), even though conservative treatment is often more severe conditions. The results of this study suggest that providing the most aggressive treatment may be an effective way to reduce mortality. Interestingly, although being female is a protective factor in this study, interaction tests revealed that untreated female patients have a greater risk than male patients, which may imply that a natural course is still a risk factor for poor prognosis in poor-grade aSAH patients. This observation suggested that as much active treatment as possible should be provided to female patients. During follow-up, 21.3% of the surviving patients were dependent on survival (mRS: 3–5) (140/655). The proportion of patients with favourable outcomes (mRS: 0-2) among all the patients was 36.8% (515/1,399), which was not significantly different from the approximately 39.7% reported at discharge (597/1,589). Wang et al.^22^ reported that the overall proportion of patients with favourable outcomes at 6 months was approximately 36.5%. Zhao et al.^24^, Robert M. Starke et al.^25^, and J Mocco et al.^26^ reported that the proportion of patients with favourable outcomes (mRS 0-2) at 12 months among total poor-grade aSAH patients ranged from 33% to 42%, which is similar to our results. Binary logistic regression analysis revealed that WFNS grade V is a significant predictor of dependency in patients with poor-grade aSAH, which is consistent with previous research^17^. Notably, middle cerebral artery aneurysm is an important risk factor for dependency in living patients, possibly because when middle cerebral artery aneurysms rupture, the haematoma near the basal ganglia causes destruction. In addition, cerebral vasospasm after haemorrhage can lead to ischaemia in the basal ganglia area, thus affecting the motor system and causing patients to have dependency status. Although this study revealed that different aneurysm treatment modalities could affect the long-term mortality of patients, they are not risk factors for dependency. This phenomenon is consistent with the conclusion of our research group that the dependency of 2-year survivors of aSAH among elderly patients is not affected by the treatment modality^27^.

This study has several limitations. First, the results of this study are subject to the well-known methodological limitations of retrospectively analysed cohorts, although we collected follow-up data prospectively. Second, this study analysed multicentre data, and differences in treatment strategies between centres may have introduced biases. However, all the centres were eligible cerebrovascular centres in China, and operators in both groups were well trained. Third, there were missing data in this study, and the database did not include complete imaging data. Finally, this study was limited to the Chinese population, and ethnicity differences should be considered.

## CONCLUSION

Our study revealed that among patients with poor-grade aSAH, 61.5% either died or became dependent. Although there has been a downward trend in recent years, the long-term mortality rate for poor-grade aSAH has reached 51%, and even among those who survived, 21.3% were dependent. The mortality rate significantly increased for patients who were WFNS Grade V, were older, had a previous history of stroke, and who received conservative treatment. Compared with surgical treatment, endovascular treatment is often more effective in reducing the mortality rate of poor-grade aSAH patients. WFNS grade V and middle cerebral artery aneurysms are independent risk factors for dependency. This study elucidates the long-term survival and dependency among poor-grade aSAH patients, contributing clinical evidence to inform prognostic evaluations and treatment strategies.

## Data Availability

The datasets used and/or analyzed during the current study are available from the corresponding author upon reasonable request.

## Acknowledgments

We would like to express our sincere gratitude to all of the patients and their relatives as well as to the professors from the Chinese Multicentre Aneurysm Database (CMAD) group.

## Supplemental Material

Table S1-S5 Figure S1-S3

